# Altered spreading of neuronal avalanches in temporal lobe epilepsy relates to cognitive performance: a resting-state hdEEG study

**DOI:** 10.1101/2022.11.16.22282392

**Authors:** Gian Marco Duma, Alberto Danieli, Giovanni Mento, Valerio Vitale, Raffaella Scotto Opipari, Viktor Jirsa, Paolo Bonanni, Pierpaolo Sorrentino

## Abstract

**Objective:** Large aperiodic bursts of activations named neuronal avalanches have been used to characterize whole-brain activity, as their presence typically relates to optimal dynamics. Epilepsy is characterized by alterations of large-scale brain network dynamics. Here, we exploited neuronal avalanches to characterize differences in the electroencephalography (EEG) basal activity, free from seizures and/or interictal spikes, between patients with temporal lobe epilepsy (TLE) and matched controls.

**Method:** We defined neuronal avalanches as starting when the z-scored source-reconstructed EEG signals crossed a specific threshold in any region and ending when all regions went back to baseline. This technique avoids data manipulation or assumptions of signal stationarity, focusing on the aperiodic, scale-free components of the signals. We computed individual avalanche transition matrices, to track the probability of avalanche spreading across any two regions, compared them between patients and controls, and related them to memory performance in patients.

**Results:** We observed a robust topography of significant edges clustering in regions functionally and structurally relevant for the TLE, such as the entorhinal cortex, the inferior parietal and fusiform area, the inferior temporal gyrus, and the anterior cingulate cortex. We detected a significant correlation between the centrality of the entorhinal cortex in the transition matrix and the long-term memory performance (delay recall Rey figure test).

**Significance:** Our results show that the propagation patterns of large-scale neuronal avalanches are altered in TLE during resting state, suggesting a potential diagnostic application in epilepsy. Furthermore, the relationship between specific patterns of propagation and memory performance supports the neurophysiological relevance of neuronal avalanches.

**Key Points:**

- Investigation of the brain dynamics during resting-state activity in patients with TLE using neuronal avalanches (i.e., large scale patterns of activation)
- We found higher transition probabilities in patients with TLE in the entorhinal cortex, inferior temporal and fusiform gyri, and anterior cingulate cortex
- We found higher eigenvector centrality of the left entorhinal cortex in the avalanche transition matrix, which was related to reduced long term memory performance
- Discussion of the potential application of the avalanche transition matrix as diagnostic tool in presurgical evaluations and epilepsy types differentiation

## 1 INTRODUCTION

Empirical evidence supports the hypothesis that the human brain displays near critical dynamics, characterized by the coexistence of large, aperiodic burst of activations (“neuronal avalanches”) and oscillatory activity (Zalesky et al., 2014). The scale-invariant properties showed by brain signalshave been associated with a dynamical regime supporting maximally efficient and flexible information processing (O’Byrne & Jerbi K, 2022).

Epilepsy is a neurological disorder characterized by recurrent, unprovoked seizures, the generation and propagation of which depends on the activation of networks of interrelated brain regions, defined as “epileptogenic network” (EN) (Bartolomei, Guye, & Wendling, 2013; Davis, Jirsa, & Schevon, 2021)

According to the first, mainly empirical conceptualization, which rose in the context of stereo-electroencephalographic presurgical assessment of focal epilepsy, the EN represents the set of brain regions involved in the primary organization of an individual’s ictal discharge, and the ultimate target of surgical treatment (Bartolomei et al., 2017). On a broader perspective, network theory is applied in the context of epilepsy as a framework to describe, with different methodologies, the complex spatio-temporal organization of the EN activity (ictal and interictal) (Bernhardt et al., 2016; Duma et al., 2021), as well as its overall clinical impact, which may include cognitive and psychosocial dysfunction (Davis, Jirsa, & Schevon, 2021, Fadaie et al., 2021,).

The critical brain hypothesis may provide a further and complementary framework to describe complex brain network dynamics, with important applications for the study of neurological disorders (Zimmern, 2020). In particular, large aperiodic bursts of activations that spread across the brain (named “neuronal avalanches”) have been observed in large-scale neurophysiological recordings (Shriki et al., 2013). More recently, avalanche transition matrices have been applied to magneto/electroencephalography (M/EEG) data, in order to encapsulate the spatio-temporal spreading of neuronal avalanches on the large scale. These studies highlighted that neuronal avalanches spread preferentially across the white-matter bundles (Sorrentino et al., 2021), and that they are drastically altered in neurodegenerative diseases (Polverino et al., 2022; Romano et al., 2022, Sorrentino et al., 2021). In the context of epilepsy, the criticality framework has been used to investigate global brain dynamics during ictal and interictal epileptic activity (Osorio et al., 2009, Meisel et al., 2012, Arviv et al., 2016) showing a deviation of the system from the critical state in both conditions.

In recent years, a significant body of research has investigated the intrinsic functional organization of brain dynamics in the resting state (RS) condition. Notably, epilepsy-related cognitive abnormalities have been linked to an altered architecture of functional brain networks at rest (Fadaie et al., 2021, Duma et al., 2022). Most of these approaches are based on the analysis of the covariation between the corresponding signals and/or spectral power, as well as the frequency-specific phase locking (i.e. synchronization). However, such techniques rely on the assumption of stationarity. In the present work, we investigated neuronal avalanches dynamics during RS in patients with temporal lobe epilepsy (TLE). We hypothesized that, even during RS, the spatial spreading of neuronal avalanches would display specific alterations in individuals with TLE compared to controls. More precisely, we predicted that large-scale neuronal avalanches, as they spread across the brain at rest, would more likely recruit regions that are known to be functionally relevant for seizure generation and propagation in TLE. Moreover, we hypothesized that abnormalities of neuronal avalanche dynamics in TLE would be associated with neuropsychological performances, which are typically altered in TLE patients. To test these hypotheses, we estimated the neuronal avalanche transition matrices from source-reconstructed EEG acquired in patients with TLE and matched healthy controls. From the resting activity, we estimated the probability of a neuronal avalanche spreading between any pair of brain regions. Comparing (edge-wise) these probabilities in the two groups allowed us to identify, in an unsupervised, data-driven fashion, the regions that displayed altered aperiodic dynamics in individuals with TLE compared to controls. Finally, we related the altered recruitment of such regions to cognitive performance.

## 2 METHOD

### 2.1 Participants

Between January 2018 and December 2020 a total of 49 patients with drug resistant TLE underwent a hdEEG recording as part of the presurgical evaluation at IRCCS E. Medea, Conegliano (Treviso), Italy. The presurgical workup included detailed clinical history and examination, neuropsychological assessment, long-term surface Video EEG monitoring, and 3T brain MRI, with PET as an adjunctive investigation in selected cases. The sample size was determined based on the subject availability of the clinical center. The inclusion criteria were: -age ≥ 18 years; -temporal seizure onset based on clinical history and ictal Video EEG recordings; -MRI evidence of epileptogenic lesion confined to the temporal lobe or negative MRI; -resting state activity recorded with high density electroencephalography (hdEEG) > = 8 min. The inclusion criteria were decided before the data analysis. A total of 37 patients, 16 left TLE, 8 right TLE, 13 bitemporal, mean age = 25.3 [SD = 19.55]; 20F) were eligible for the study.

The mean number of antiseizure medications (ASM) per person was equal to 2.05 with a range going from a minimum of 0 to a maximum of 5 ASM. Specifically, among the patients with TLE 3 had no medication, 9 were in monotherapy and all the rest of participants were in polytherapy. A description of patients’ demographic and clinical characteristics is given in Table 1.

**Table 1.**
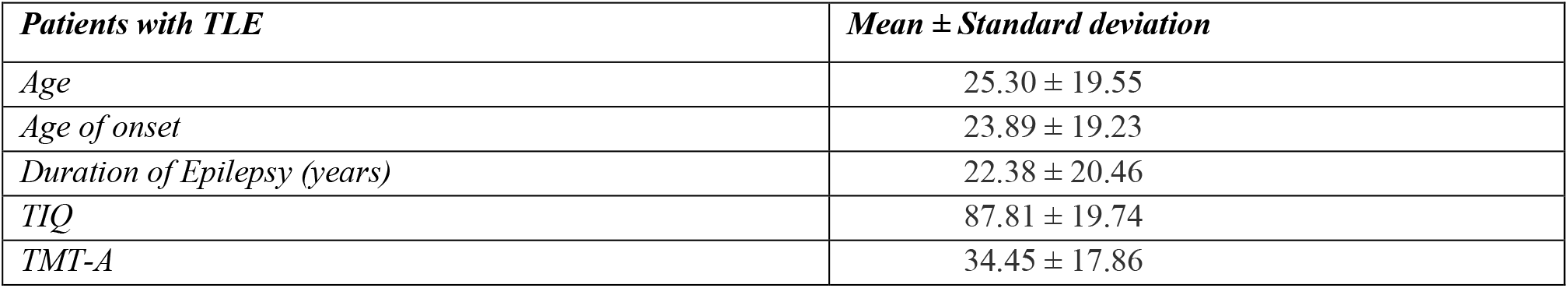
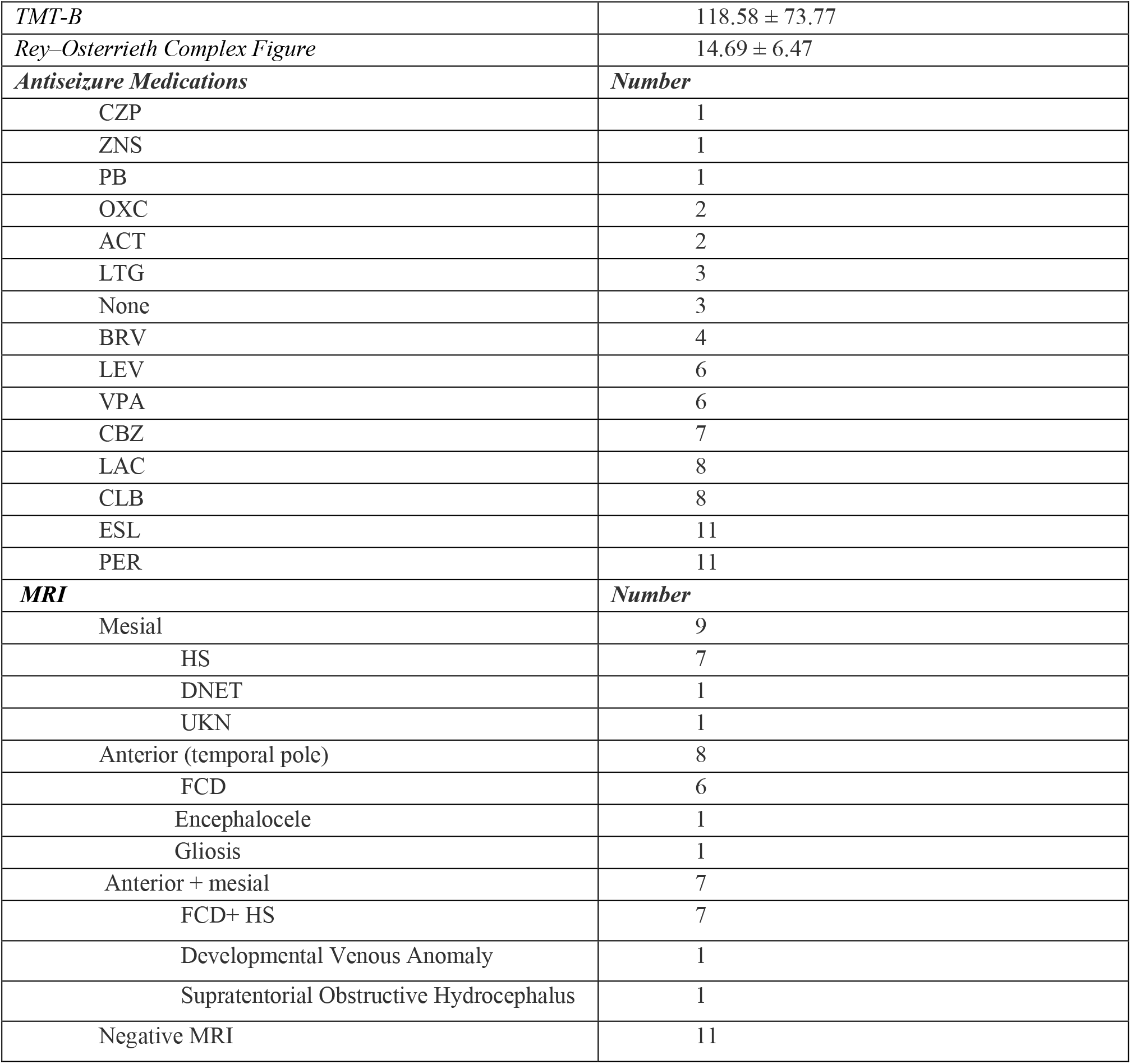
The table describes the demographic and clinical characteristics of the patients with temporal lobe epilepsy, along with the scores of the neuropsychological tests. MRI abnormalities are reported by sublobar localization. The continuous variables are reported as mean ± standard deviation. Antiseizure Medication abbreviations: NFT = no pharmacological treatment, CBZ = carbamazepine, CLB = clobazam, CZP = clonazepam, LEV = levetiracetam, LTG = lamotrigine, PER = perampanel, VPA= valproic acid, ESL= eslicarbazepine, LAC = lacosamide, OXB = oxcarbazepine, BRV = bivaracetam, ACT= acetazolamide, ZNS = zonisamide, PB = phenobarbital. Abbreviation of the identified anomalies on the magnetic resonance imaging: FCD= focal cortical dysplasia, HS=hippocampal sclerosis, DNET = dysembryoplastic neuroepithelial tumors, UKN = unknown.

| *Patients with TLE* | *Mean ± Standard deviation* |
| --- | --- |
| *Age* | 25.30 ± 19.55 |
| *Age of onset* | 23.89 ± 19.23 |
| *Duration of Epilepsy (years)* | 22.38 ± 20.46 |
| *TIQ* | 87.81 ± 19.74 |
| *TMT-A* | 34.45 ± 17.86 |
| *TMT-B* | 118.58 ± 73.77 |
| *Rey–Osterrieth Complex Figure* | 14.69 ± 6.47 |
| ***Antiseizure Medications*** | ***Number*** |
| CZP | 1 |
| ZNS | 1 |
| PB | 1 |
| OXC | 2 |
| ACT | 2 |
| LTG | 3 |
| None | 3 |
| BRV | 4 |
| LEV | 6 |
| VPA | 6 |
| CBZ | 7 |
| LAC | 8 |
| CLB | 8 |
| ESL | 11 |
| PER | 11 |
| ***MRI*** | ***Number*** |
| Mesial | 9 |
| HS | 7 |
| DNET | 1 |
| UKN | 1 |
| Anterior (temporal pole) | 8 |
| FCD | 6 |
| Encephalocele | 1 |
| Gliosis | 1 |
| Anterior + mesial | 7 |
| FCD+ HS | 7 |
| Developmental Venous Anomaly | 1 |
| Supratentorial Obstructive Hydrocephalus | 1 |
| Negative MRI | 11 |

The control group was composed of 35 healthy participants with no history of neurological or psychiatric disorders (mean age = 35.10 [SD = 8.24]; 28 F). The study was conducted according to the principles expressed in the Declaration of Helsinki and approved by the local ethical committee.

### 2.2 Neuropsychological scores

All the patients with TLE underwent a neuropsychological evaluation focusing on memory, attention/executive functions and intelligence. Specifically, long term memory was investigated with the delay recall of the Rey–Osterrieth Complex Figure Test (ROCFT) (Caffarra et al., 2000). Attention and executive functions were evaluated with the Trail Making Test (TMT) (Giovagnoli et al., 1996). Specifically, we included in the analysis both the part A and B as a measure of motor speed and shifting capabilities, respectively. Finally, we used the total IQ (TIQ) of the WAIS-IV scale (Orsini & Pezzuti, 2013) as a global intelligence measure. However, the evaluation was not completed by every subject. Therefore, we reduced the sample size to 27 patients for the analyses of the correlation of the neuropsychological functioning with the neural activity Summary statistics for each test score in the clinical population is provided in Table1.

### 2.3 Resting State EEG recording

The hdEEG recordings were obtained using a 128-channel Micromed system referenced to the vertex. Data was sampled at 1,024 Hz and the impedance was kept below 5kΩ for each sensor. For each participant we recorded 10 minutes of closed-eyes resting state while comfortably sitting on a chair in a silent room.

### 2.4 EEG pre-processing

Signal preprocessing was performed through EEGLAB 14.1.2b (Delorme & Makeig, 2004). The continuous EEG signal was first downsampled at 256 Hz and then bandpass-filtered (0.1 to 45 Hz) using a Hamming windowed sinc finite impulse response filter. The signal was visually inspected to identify interictal epileptiform discharges (IEDs) by GMD, AD and PB and then segmented into 2 sec epochs. Epochs containing IEDs activity were successively removed. The epoched data run through an automated cleaning algorithm using the TBT plugin implemented in EEGLAB. This algorithm identified the channels that exceeded a differential average amplitude of 250μV and marked those channels for rejection. Channels that were marked as bad on more than 30% of all epochs were excluded. Additionally, epochs having more than 10 bad channels were excluded. We automatically detected possible flat channels with the Trimoutlier EEGLAB plug in with the lower bound of 1μV. We rejected an average of 16.43 ± 10.91 (SD) epochs due to spikes and 4.45 ± 2.91 (SD) due to artifacts. The previous mentioned preprocessing analysis pipeline has been applied by our group in previous studies investigating both task-related and resting state EEG activity (Duma et al., 2022; Duma, Di Bono, Mento, 2021). The data cleaning was performed using independent component analysis (Stone, 2002), implemented via the Infomax algorithm (Bell & Sejnowski, 1995) in EEGLAB. The resulting 40 independent components were visually inspected (both the topography and the time-series), and those related to eye blinks, eye movements, muscle or cardiac artifacts were discarded. The remaining components were then back-projected to the electrode space. Finally, bad channels were reconstructed with the spherical spline interpolation method (Perrin et al., 1989). The data were then re-referenced to the average of all electrodes. At the end of the data preprocessing, each subject had at least 8 minutes of artifact-free signal.

### 2.5 Cortical Source modelling

We used the individual anatomy MRI in order to generate individualized head models for the patients with TLE. The anatomical MRI for source imaging consisted of a T1 1 mm isotropic 3D acquisition (256 × 256 × 256 matrix). For 3 patients and for the control group we used the MNI-ICBM152 default anatomy (Evans et al., 2012) from Brainstorm (Tadel et al., 2011) since the 3D T1 MRI sequences were not available. The MRI was segmented in skin, skull and gray matter using the segmentation pipeline of the Computational Anatomy Toolbox (CAT12; Christian Gaser 2018, http://www.neuro.uni-jena.de/cat/). The resulting individual surfaces were then imported in Brainstorm, where three individual surfaces adapted using the Boundary Element Models (BEM) were reconstructed (inner skull, outer skull and head) and the cortical mesh was downsampled at 15,002 vertices. Co-registration of EEG electrodes was performed using Brainstorm by projecting the EEG sensors positions on the head surface according to the fiducial points of the individual MRI. We applied slight manual correction to better orient the EEG cap on the individual anatomy, and then we projected the electrodes on the individual head surface using Brainstorm. We then derived an EEG forward model using the 3-shell BEM model (conductivity: 0.33, 0.165, 0.33 S/m; ratio: 1/20) (Gonçalves et al., 2003) estimated using the OpenMEEG method implemented in Brainstorm (Gramfort et al., 2010), which provides a realistic head model. Finally, we used the weighted minimum norm estimation (Baillet et al., 2001) as the inverse model, with the default parameter settings provided in Brainstorm.

### 2.6 Avalanche estimation

We extracted the activity of a total of 68 regions of interest (ROIs) from the Desikan-Killiany atlas (Desikan et al., 2008). The ROIs time series was obtained by averaging the activity across the vertices composing each ROI. To study the dynamics of brain activity, we estimated “neuronal avalanches” from the source-reconstructed ROI time series. Firstly, the time series of each ROI was binarized by calculating the z-score across time and then identifying positive and negative excursions beyond a threshold.

A neuronal avalanche begins when, in a sequence of contiguous time bins, at least one ROI is active (i.e. above the threshold of |z-score| >3), and it ends when all ROIs are inactive (Beggs & Plenz, 2003; Shriki et al., 2013). Alternative z-score thresholds (i.e., 2.9 and 3.1) were tested. The total number of active ROIs in an avalanche corresponds to its size. These analyses require the time series to be binned. This is done to ensure that one is capturing critical dynamics, if present. To estimate the suitable time bin length, for each subject, each neuronal avalanches, and each time bin duration, the branching parameter σ was estimated (Haldeman and Beggs, 2005). In fact, systems operating at criticality typically display a branching ratio ∼1. The branching ratio is calculated as the geometrically averaged (over all the time bins) ratio of the number of events (activations) between the subsequent time bin (descendants) and that in the current time bin (ancestors) and then averaging it over all the avalanches (Bak et al., 1987). More specifically:

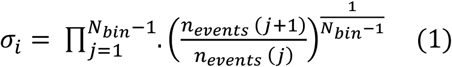

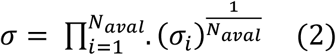

where σ _*i*_ is the branching parameter of the *i*^*th*^ avalanche in the dataset, *N*_*bin*_ is the total amount of bins in the *i*^*th*^ avalanche, *n*_*events*_ *(j)* is the total number of events active in the jth bin, and *N*_*aval*_ is the total number of avalanche in the dataset. In our analyses the branching ratio was 1 for bin = 2 (8 ms).

### 2.7 Transition matrices

Avalanches shorter than 2 time bins (∼8 ms) were excluded from further analyses. However, the analyses were repeated also including only avalanches longer than 4 time bins (∼16 ms), in order to focus on rarer events (the sizes of the neuronal avalanches have a fat-tailed distribution) that are highly unlikely to be due to noise, and the results were unchanged (Marshall et al., 2016) (see Supplementary Figure 2). An avalanche-specific transition matrix (TM) was calculated, where element in position (*i, j*) represented the probability that region j was active at time *t + σ*, given that region *i* was active at time ***t***, where σ ∼ 8 ms (see also Fig.1). The TMs were averaged per participant, and then per group, and finally symmetrized. The introduction of a time lag makes it unlikely that our results can be explained trivially by volume conduction (i.e., the fact that multiple sources are detected simultaneously by multiple sensors, generating spurious zero-lag correlations in the recorded signals). For instance, for a binning of 2, as the avalanches proceed in time, the successive regions that are recruited do so after roughly 8 ms. Hence, activations occurring simultaneously do not contribute to the estimate of the TM.

### 2.8 Statistics

For each group, we computed the difference in the probability of a perturbation running across a given edge in patients and controls. To statistically validate the difference in the transition matrices across groups (patients with epilepsy vs. control group), we randomly shuffled the labels of the TMs across groups (i.e. each subject-specific transition matrix was randomly allocated to either the patients or the controls group). We performed this procedure 10,000 times, obtaining, for each edge, the distribution of the differences given the null-hypothesis that the TMs would not capture any difference between the two groups. We used the null distribution to obtain a statistical significance for each edge. We applied False Discovery Rate (FDR) correction for multiple comparisons across edges (Benjamini & Hochberg, 1995). Following this procedure, we obtained a matrix with the edges whose probabilities significantly differed between the two groups. Successively, we checked if the significantly different edges would cluster over specific brain regions. To this end, we computed the expected number of significant edges incident on any region, given a random distribution (with a comparable density), and selected those regions with an above-chance number of significant edges clustered upon them. Statistics were again corrected using FDR, this time across regions.

To check the consistency of the results we performed statistical analyses also for the z-score threshold of 2.9 and 3.1, and a different time binning including only avalanches longer than 4 time bins (16 ms). See figure 1 for the graphical representation of the analysis pipeline.

**FIGURE 1.**
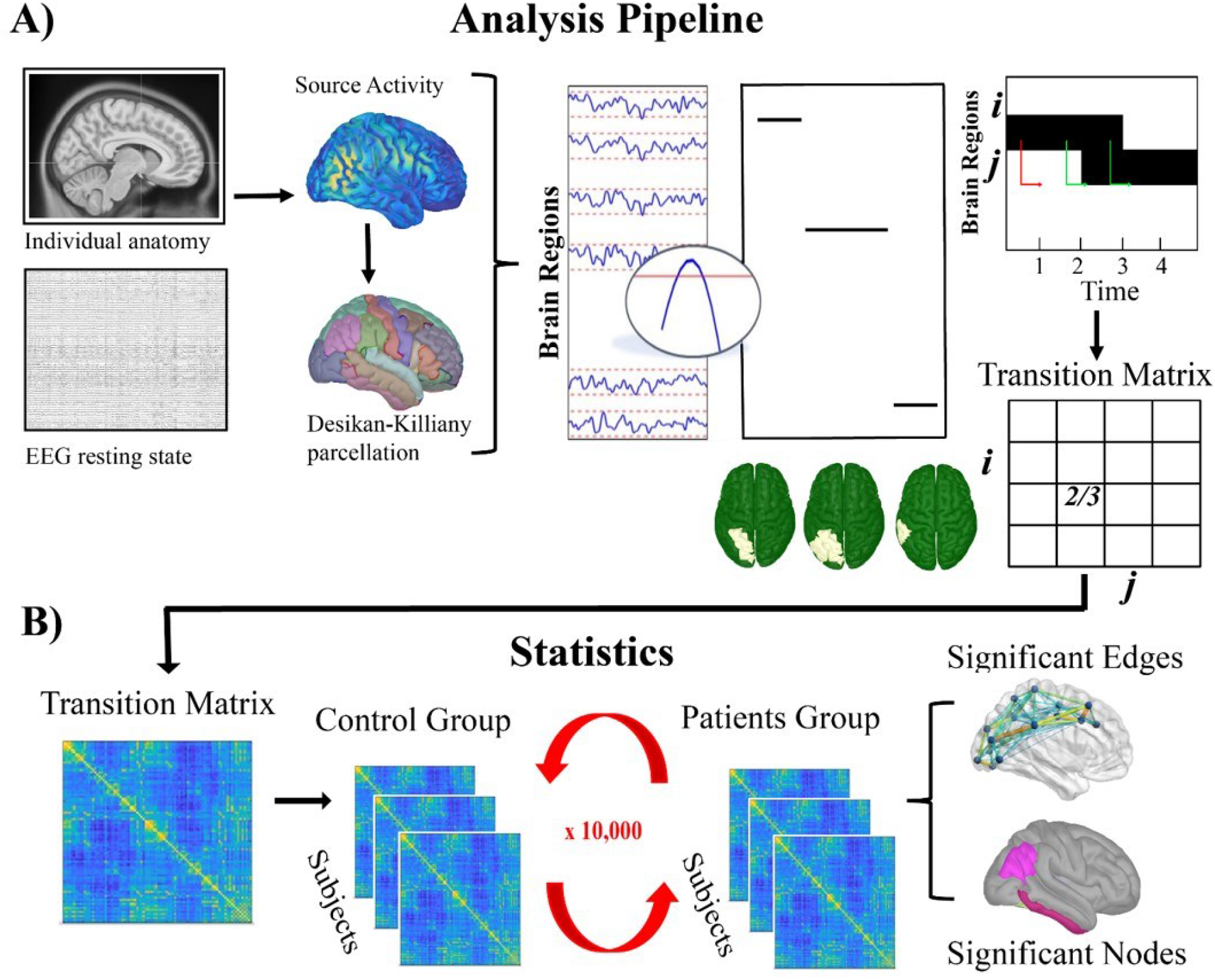
Graphical representation of analysis pipeline. The panel A of the figure represents the source reconstruction of the signal from resting-state EEG, and the following generation of the avalanche transition matrix after the source activity thresholding. Panel B illustrates the statistical procedure of the 10,000 permutation to extract significant edges and nodes.

#### 2.8.1 Correlations with the neuropsychological functioning

Finally, we investigated the relationship between the probability of an avalanche recruiting a specific brain region and the neuropsychological functioning. At first we extracted centrality measures for each node in the ATMs, namely the eigenvector centrality, using the Brain Connectivity Toolbox (Rubinov & Sporns, 2010). Successively, we checked for the normality of data distribution using the Shapiro-Wilk test. We observed that the data graph indices were not normally distributed in the patients group (p = .046). For this reason, we applied Spearman’s correlation between graph theory indices of the statistically significant nodes and the neuropsychological measures, separately for each graph measure. We then applied FDR correction across tests. Considering the relatively small sample size, in order to reduce the effect of the extreme value in the correlation computation, we applied winsorized robust correlations using the WRS2 package in R. In this case we specifically targeted the significant results coming from the FDR-corrected correlation analysis.

## 3 RESULTS

We used the spatio-temporal spreading of large aperiodic bursts of activations as a proxy for communications between pairs of regions. Within this framework, large-scale, higher-order perturbations are considered to mediate the interactions between brain regions. We tested for differences between the two groups (i.e. patients with TLE and healthy controls) in the probabilities of any such perturbation to propagate across two brain regions. To this end, we built an avalanche transition matrix (ATM) for each subject, containing regions in rows and columns, and the probability that region *j* would activate at time (*t*+1), given that region *i* was active at time t, as the *ij*^*th*^ entry. The differences in the probability of being sequentially recruited by an avalanche was used to track (subject- and edge-wise) the difference on the spatial propagation of the perturbations across the two groups. To validate the observed differences, for each subject and each edge, we built a null-model randomizing the labels (i.e. patients or healthy control) 10,000 times, in order to obtain a null distribution of the differences expected by chance. These distributions were used to spot, individually, the edges that differed between the two conditions above chance level. We applied FDR to correct for multiple comparisons across edges.

As a first result we observed that the significant edges across groups were preferentially hinging the temporo-frontal areas, suggesting that patients with TLE show increased transition probability in these areas (see Fig. 2B).

**FIGURE 2.**
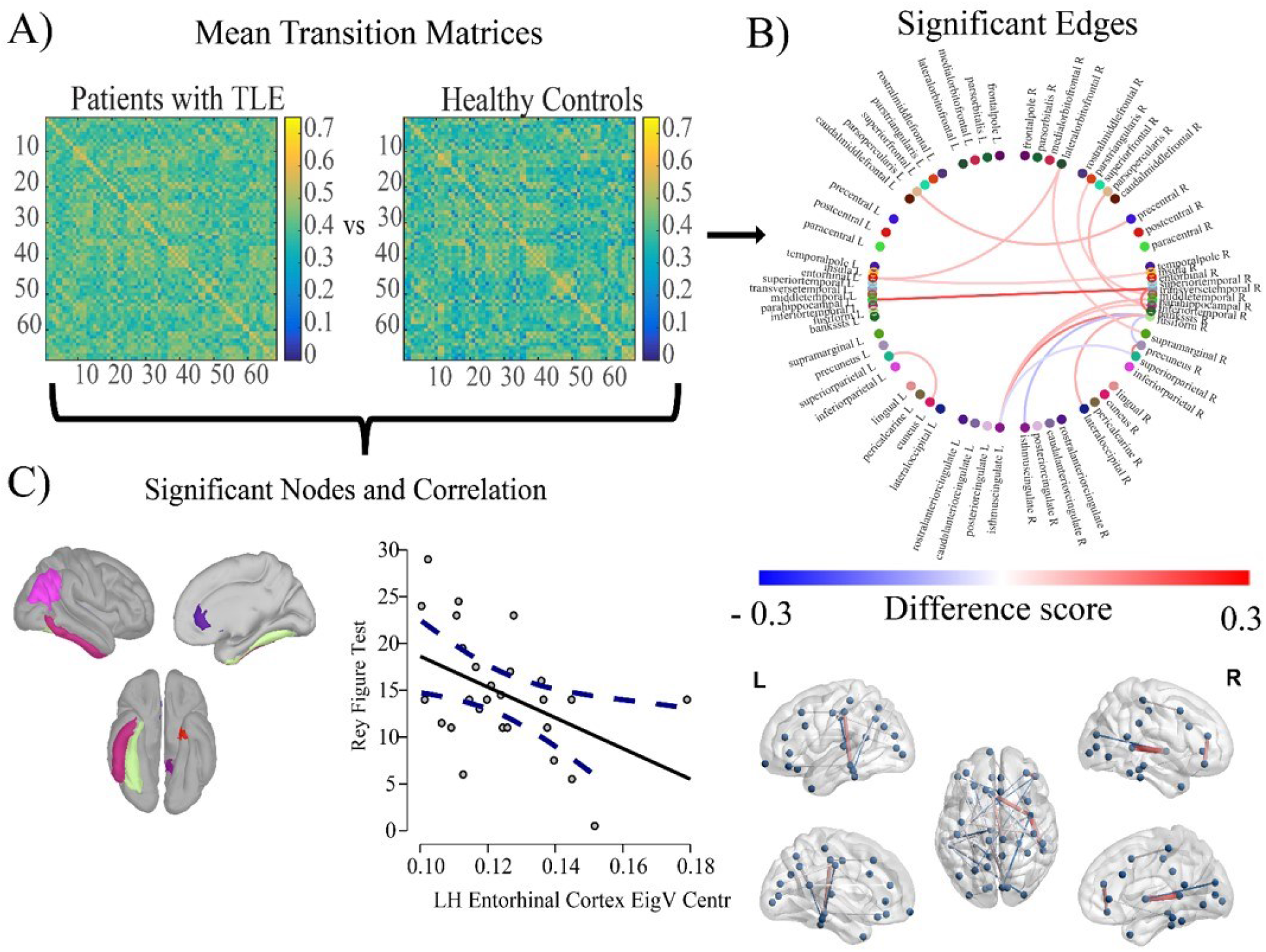
Results of the analyses. Panel A represents the group average of the avalanche transition probability matrices. Panel B (upper and lower parts) illustrates the significant edges differing between patients with TLE and the control group. Finally, panel C indicates the significant nodes differing across groups, and the correlation of the left entorhinal cortex with the long term memory performance (delay recall Rey-Osterrieth Figure Test).

We then evaluated whether the significantly different edges would significantly cluster over specific brain regions. To this end, we computed the expected number of significant edges incident on any region, given a random network (with a comparable density as the observed ones), and selected those regions with an above-chance number of significantly different edges clustered upon them. Statistics were again corrected using FDR, this time across regions. These “reliably different’ edges cluster preferentially upon the temporal and frontal regions. In particular, the left entorhinal cortex, the inferior parietal lobe, the left isthmus of the cingulate cortex, the right fusiform area, the right inferior and transversal temporal cortex, and the right rostral anterior cingulate cortex all showed to have significantly-different edges clustered upon them (p <0.001, FDR corrected).

These results were consistent across multiple z-score thresholds (2.9 and 3.1) as well as different time binnings (avalanche longer than 16 ms) (see Supplementary Fig 1 and 2).

Finally, we computed the eigenvector centrality (EC) in the ATM of each patient and, for the regions where different edges clustered, we related the individual EC to the neuropsychological scores. We observed a significant correlation (*r* = -.437; *p*_*adj*_ = .045) between the eigenvector centrality of the left entorhinal cortex and the score of the delayed recall of the Rey-Osterrieth complex figure in the group of patients with TLE. The results were robust to the outliers, as checked by the winsorized robust correlation (*r* = -.467; *p* = .013).

To test the consistency of the results, we tested the above mentioned correlation across multiple z-score thresholds. Both with the z-score threshold of 2.9 and 3.1, and avalanche longer than 4 time bins, and we observed a significant correlation between the eigenvector centrality of the left entorhinal cortex and the score of the delayed recall of the Rey-Osterrieth complex figure (see Supplementary Table 1).

## 4 DISCUSSION

In the present work we investigated the possible differences in the spontaneous, large-scale aperiodic activity occurring at rest in patients with TLE as compared to healthy controls.

We found that fast bursts of aperiodic activations (“neuronal avalanches”) during resting-state are more likely to spontaneously spread across temporal areas in patients as compared to controls. We were able to provide such mapping starting from resting state, identifying areas that are typically involved in critical events in TLE patients, and also display a (presumably) pathological role on the large-scale functional networks, regardless of the presence of epileptiform activity. By utilizing the avalanche transition matrices, i.e. a mathematical tool that accommodates the non-linearities of the large-scale brain dynamics and the corresponding multimodal dynamics that they generate, we successfully identified the anomalous spreading of activity in patients with TLE from resting-state EEG data. The investigation of brain dynamics through the criticality framework represents a useful tool to describe the mechanisms underpinning the evolution of brain activity in terms of the aperiodic dynamics. However, criticality and neural avalanche have been mostly applied to identify epileptogenic areas (Worrell et al., 2002; Witton et al., 2019) or characterizing brain dynamics during seizures (Osorio et al., 2009, 2010; Meisel et al., 2012). Importantly, our work is focused on the basal brain activity, assessing the intrinsic network organization of the brain with epilepsy when epileptiform activity is not occurring. Relevantly, while derived from the framework of criticality, the ATMs allowed us to move away from the traditional studies of criticality which focused on the global properties of avalanches. In fact, we were able to evaluate the internal dynamics of avalanches, which may provide more useful information on the specific brain regions involved in epilepsy.

This line of thinking builds upon converging evidence indicating that the brain alternates segregated and integrated states (Cohen & D’Esposito, 2016; Fukushima & Sporns, 2020). As such, meaningful communication among regions on the large-scale would be intermittent, and might be best understood and measured in terms of aperiodic perturbations. We reasoned that, if avalanches convey interactions occurring between regions, then their spreading should also be modified between groups, if there is a different communicative basal pattern. Our results identified, in an unsupervised manner, a number of functional links (i.e. edges) that are more likely to be dynamically recruited in patients with TLE. The edges are mainly incident on temporal and fronto-central regions. These results align with the evidence of altered functional connectivity of temporal and frontal areas in TLE (Maccotta et al., 2013; Qin et al., 2020). We observed that the significantly different edges predominantly hinged upon temporal regions such as the left entorhinal cortex, the fusiform area, as well as the superior temporal gyrus. These regions are known to be involved in TLE. In particular, the entorhinal cortex is considered a key player in seizure initiation in TLE (Rutecki et al., 1989; Vismer et al., 2015).

Furthermore, individuals with TLE frequently display altered functioning, often associated with a hyper-activation, of the network involved in face recognition, including the fusiform area (Guedj et al., 2011; Riley et al., 2015), which we also find altered Moreover, our results highlighted the involvement of the anterior cingulate cortex (ACC), which has been previously reported in TLE. In fact, evoked responses in the cingulate cortex have been detected after hippocampal stimulation in patients with TLE (Kubota et al., 2013). Additionally, more recent evidence highlighted altered resting state functional connectivity of the ACC in medial TLE (Jo et al., 2019

Importantly, the alterations in the avalanche spreading that we observed in TLE correlated with cognitive functioning. Specifically, we observed that the more the entorhinal cortex was recruited by the transient bursts (i.e., higher centrality in the ATM), the worse the cognitive performance in the long term memory tasks (measured by the Rey figure test). This finding is particularly interesting, as it endows the spreading of activations in the data to a functional meaning, since it relates to a cognitive process that has been long known to be impaired in patients with TLE. In addition to this, it may be worth noticing that the vast majority of large-scale brain imaging studies has investigated the interactions among brain regions exploiting the statistical dependencies between the corresponding signals. With regard to epilepsy, one of the main focuses has been to characterize the behavior of brain networks during seizures or interictal epileptiform discharges (IED), since they can provide most relevant information in relation to diagnosis and also to treatment selection and planning. In contrast, here we exploited the spontaneous dynamics embedded in the simple raw signal, derived from source activation maps of the resting activity, purposely disregarding epileptiform activity. Strikingly, disregarding most of the signals and focusing on the aperiodic transients has allowed us to apply a remarkably simple pipeline, which avoids heavy filtering, assumptions of stationarity, and is applied in the time-domain. In fact, by thresholding the z-scored signals, temporal regions naturally emerged as key players in the resting-state dynamics, beyond ictal or IED. However, it is important to note that we may have underestimated the true frequency of IED, since they may not be evident on surface EEG.

At the same time, it remains difficult to provide an unambiguous mechanistic interpretation of our findings. It might be that a potential alteration of the regional excitation/inhibition (E/I) balance could facilitate the recruitment of a given brain region by the ongoing transient large-scale perturbations. In the same line of thinking, Burrows and colleagues (Burrows et al., 2021) observed that manipulating the E/I balance in cellular in vivo neural networks caused a shift of the operational regime away from criticality which was linked, in turn, to the emergence of epileptic activity. Beyond the neuroscientific implications, our methodology might directly help the patients’ management. In particular, the present methodology might serve as a non-invasive and cost-effective tool exploiting basal activity in order to identify specific profiles of functionally altered regions in different forms of epilepsy. Moreover, it can be used in complex diagnostic processes, such as those in which scalp EEG monitoring failed in recording seizure activity. In these scenarios, our methodology allows the identification of functionally altered zone that may be the target for additional investigation (e.g., intracranial recording). However, longitudinal studies are required to validate such findings in relation to clinical outcomes after surgical resection.

## 5 CONCLUSIONS

Our study evaluated brain dynamics during resting-state activity in TLE as compared to healthy controls utilizing the framework of neuronal avalanche. We found specific alterations of the transition probability in the entorhinal cortex, the inferior temporal and fusiform gyri, and the anterior cingulate cortex. Importantly, higher eigenvector centrality of the left entorhinal cortex was related to lower long-term memory performance. The present methodology might serve as a potential diagnostic tool to identify functionally altered regions, which might in turn aid the diagnostic process.

## Data Availability

Given patients' sensitive information, the databases and images used during the current study are not publicly archived. This restriction is required by the conditions of the ethical approval. Data can be shared upon request to the authors, after the approval by the local ethics committee and the completion of the data sharing agreement. Legal copyright restrictions prevent public archiving of the Rey Osterrieth Complex Figure Test, Trail Making Test, and The Wechsler Adult Intelligence Scale (WAIS IV) which can be obtained from the copyright holders in the cited references. The code will be made freely available on GitHub.

## ACKNOWLEDGMENTS

This work was supported by a 2019 “5XMille” funds for biomedical research of The Italian Health Ministry to PB and by the European Union’s Horizon 2020 research and 569 innovation program under grant agreement No. 945539 (SGA3).

## CONFLICT OF INTERESTS

None of the authors has any conflict of interest to disclose.

## ETHICAL PUBLICATION STATEMENT

We confirm that we have read the Journal’s position on issues involved in ethical publication and affirm that this report is consistent with those guidelines

## AUTHOR CONTRIBUTIONS

Conceptualization: Gian Marco Duma, Pierpaolo Sorrentino. Methodology: Gian Marco Duma, Pierpaolo Sorrentino. Writing–original draft preparation: Gian Marco Duma, Pierpaolo Sorrentino, Alberto Danieli, Giovanni Mento, Viktor Jirsa, Paolo Bonanni. Formal analysis: Gian Marco Duma, Pierpaolo Sorrentino. Data curation: Gian Marco Duma, Valerio Vitale, Raffaella Scotto Opipari. Investigation: Gian Marco Duma, Pierpaolo Sorrentino, Alberto Danieli, Giovanni Mento, Viktor Jirsa, Paolo Bonanni. Visualization: Gian Marco Duma, Pierpaolo Sorrentino. Supervision: Viktor Jirsa, Alberto Danieli, Paolo Bonanni, Pierpaolo Sorrentino, Funding acquisition: Paolo Bonanni

## Supplementary Materials

To check for the consistency of the results, we used different z-score thresholds, namely 2.9 and 3.1, in the avalanche detection, as well as a different time binning (i.e., avalanches > 4 time bins). Successively, we computed transition matrices and we performed statistics as described in the method section of the manuscript. Additionally, we also performed correlation between left entorhinal cortex eigenvector centrality of transition matrix and the delayed recall of Rey Figure test. Here below the table with the additional results of the correlation and the figures with the significant edges and nodes. As it is possible to notice the main results of the manuscript are confirmed even by sweeping across different z-score thresholds. Across the thresholds in fact we observe stables egged clustering in the following nodes: entorhinal cortex, inferior parietal, fusiform area, inferior temporal gyrus and anterior cingulate cortex

| **Z-score threshold** | **Spearman correlation** | **p-val** |
| --- | --- | --- |
| **2.9** | - .386 | .046 |
| **3.1** | -.397 | .040 |
| **Time bin** | **Spearman correlation** | **p-val** |
| **> 4** | -.422 | .028 |

**SUPPLEMENTARY FIGURE 1.**
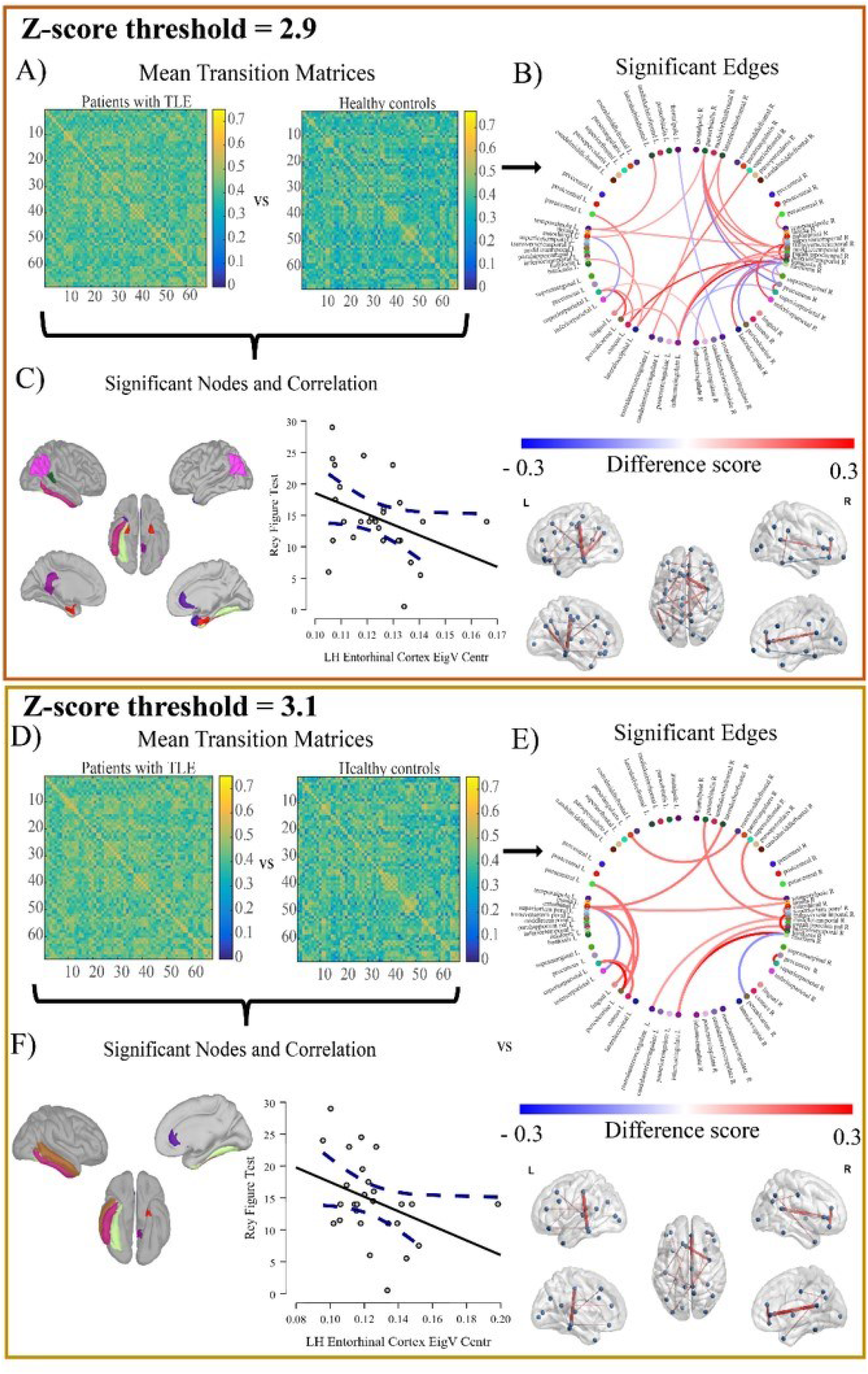
Multiple z-score threshold results. The present figure displays the results with multiple z-score thresholds (2.9; 3.1). Panel A and D represents the group average of the avalanche transition probability matrices. Panel B and E (upper and lower parts) illustrates the significant edges differing between patients with TLE and the control group. Finally, panel C and F indicate the significant nodes differing across groups and the correlation of the left entorhinal cortex with the long term memory performance (delay recall Rey-Osterrieth Figure Test).

**SUPPLEMENTARY FIGURE 2.**
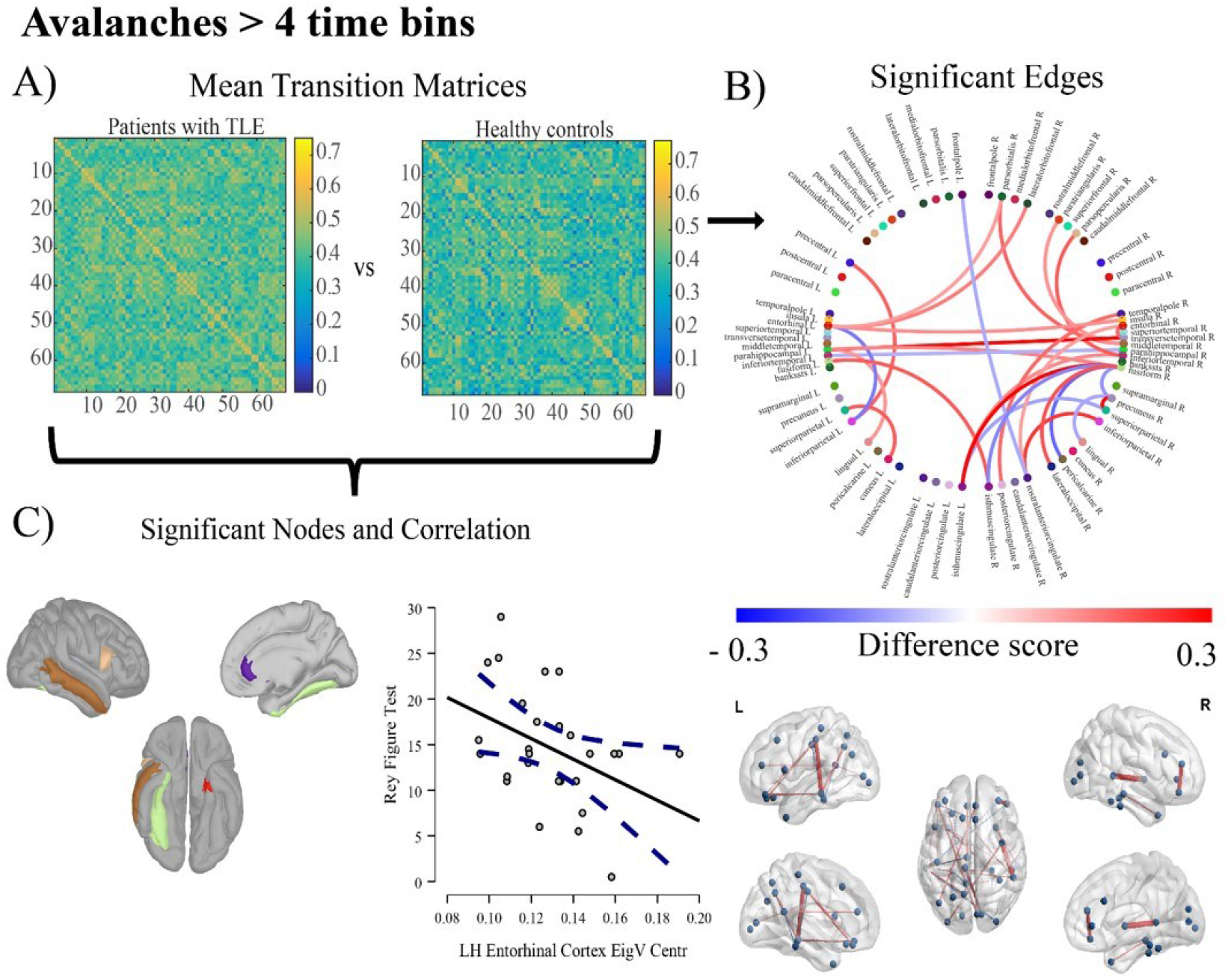
Results with avalanches larger than 4 time bins. The panel A of the present figure represents the group average of the avalanche transition probability matrices. Panel B (upper and lower parts) illustrates the significant edges differing between patients with TLE and the control group. Finally, panel C indicates the significant nodes differing across groups and the correlation of the left entorhinal cortex with the long term memory performance (delay recall Rey-Osterrieth Figure Test).

## Notes

### Competing Interest Statement

The authors have declared no competing interest.

### Author Declarations

Comitato Etico per la Sperimentazione Clinica (CESC) delle province di Treviso e Belluno, istituito con deliberazione del Direttore Generale n. 63 del 26/01/2017.

